## Supplementary material for "Body mass index and all-cause mortality in HUNT and UK Biobank studies: revised non-linear Mendelian randomization analyses"

We here perform a simulation study investigating whether adjusting genetic associations for factors that influence selection reduces selection bias within strata in a non-linear Mendelian randomization framework. We generate simulated data for 1,000,000 individuals on a genetic variant *G*, an exposure *X*, an outcome *Y*, and competing risk factors *U* and *V*. For simplicity, all variables are modelled as continuous and normally distributed.

We consider two scenarios. In each scenario, the genetic variant is a valid instrumental variable in the population as a whole, and there is no true effect of the exposure on the outcome. This implies that the genetic variant should not be associated with the outcome. We estimate the association between the genetic variant and the outcome in a selected subset of the population.

In scenario 1, the data-generating model is:

$$G_{i} \sim Normal\left( 0,1 \right)$$

$$U_{i} \sim Normal\left( 0,1 \right)$$

$$V_{i} \sim Normal\left( 0,1 \right)$$

$$X_{i} \sim Normal\left( 0.2 G_{i}+0.4 U_{i}+0.4V_{i}, 1 \right)$$

$$Y_{i} \sim Normal(-0.5 U_{i}+0.2 V_{i}, 1)$$

Participants (indexed by the *i* subscript) are selected into the sample with probability $expit(-2+U_{i}+X_{i})$, where $expit\left( x \right)=\frac{exp(x)}{1+exp(x)}$ is the inverse of the logistic (logit) function. Here *U* and *V* are both confounders of the relationship between the exposure and outcome, and selection depends on both *U* and *X*. We compare genetic associations with the outcome with and without adjustment for *U*.

In scenario 2, the data-generating model is:

$$G_{i} \sim Normal\left( 0,1 \right)$$

$$V_{i} \sim Normal\left( 0,1 \right)$$

$$X_{i} \sim Normal\left( 0.2 G_{i}+0.4 V_{i}, 1 \right)$$

$$U_{i} \sim Normal\left( 0.3 X_{i},1 \right)$$

$$Y_{i} \sim Normal(-0.5 U_{i}+0.2 V_{i}, 1)$$

Participants are selected into the sample with probability $expit(-2+U_{i})$. Here *V* is a confounder of the relationship between the exposure and outcome, and *U* is a downstream effect of the exposure *X*, and hence *U* is a collider for *G* and *V*. Selection depends on *U* only. Again, we compare genetic associations with the outcome with and without adjustment for *U*.

In both cases, all normal distributions are independent. We simulate 1000 datasets for each scenario, and compare the mean estimate of the genetic association with the outcome and its empirical Type 1 error rate at a 5% significance level (that is, the proportion of 95% confidence intervals excluding the null). Around 15-20% of the 1,000,000 participants are selected in each simulated dataset.

In scenario 1, the mean association of the genetic variant with the outcome without adjustment for *U* was 0.014 (mean standard error 0.002), and the empirical Type 1 error rate was 100.0%. With adjustment for *U*, the mean association was -0.002 (mean standard error 0.002), and the empirical Type 1 error rate was 17.1%.

In scenario 2, the mean association of the genetic variant with the outcome without adjustment for *U* was -0.025 (mean standard error 0.003), and the empirical Type 1 error rate was 100.0%. With adjustment for *U*, the mean association was -0.001 (mean standard error 0.003), and the empirical Type 1 error rate was 7.8%.

In both cases, bias reduced considerably on adjustment for the factor influencing selection. In scenario 1, mean bias did not reduce to zero, as selection also depends on the exposure. We note that if selection did not additionally depend on the exposure, there would not be selection bias, as *U* is a confounder not a collider. However, even in this case with imperfect adjustment, the reduction in bias was substantial. In scenario 2, mean bias was indistinguishable from zero in the limited number of simulated datasets considered. Type 1 error rate was slightly higher than the expected 5% rate; this is likely due to the non-linear relationship between the selection factor *U* and the probability of selection.

This simulation study is a deliberately designed to be relatively simple and limited in scope, as we do not believe that any simulation study would be able to prove conclusively that covariate adjustment eliminates selection bias in all cases. Indeed, we do not make that claim; rather we claim that adjustment for the key predictors of selection is likely to ameliorate bias due to selection. There will always be intrinsic uncertainty in practice, as we do not know all the factors that influence selection, and we would be cautious to adjust for too many variables in a Mendelian randomization analysis, particularly for variables that could be mediators on the causal pathway from the exposure to the outcome, or could be colliders (common effects) of the exposure and outcome. As age and sex cannot logically be caused by the exposure, they can be safely adjusted for without risk of these variables being mediators or colliders [S1]. Hence, if age and sex are strong predictors of selection into the sample, then adjustment for age and sex should reduce selection bias in Mendelian randomization investigations.

We believe that any residual bias from selection is likely to be low, in line with previous simulation studies that have shown selection effects need to be substantial in magnitude to have non-negligible biasing effects on Mendelian randomization estimates [S2]. However, we acknowledge that this imposes an additional level of uncertainty on the results of a non-linear Mendelian randomization analysis, on top of the existing uncertainty in the Mendelian randomization paradigm due to the validity of the instrumental variable assumptions, and other factors. All methods for making causal inferences from observational data rely on untestable assumptions, and findings should be interpreted with appropriate caution.

**Supplementary Table 1:** Genetic variants and associations with BMI based on European sex-combined analysis from GIANT ^23^. Beta-coefficents are in standard deviation units.

| SNP | Effect allele^*^ | Other allele | Beta-coefficient | Standard error of beta | Number of subjects | *P*-value |
| --- | --- | --- | --- | --- | --- | --- |
| rs1558902 | A | T | 0.082 | 0.003 | 320 073 | 7.51E-153 |
| rs6567160 | C | T | 0.056 | 0.004 | 321 958 | 3.93E-53 |
| rs10938397 | G | A | 0.040 | 0.003 | 320 955 | 3.21E-38 |
| rs543874 | G | A | 0.048 | 0.004 | 322 008 | 2.62E-35 |
| rs2207139 | G | A | 0.045 | 0.004 | 322 019 | 4.13E-29 |
| rs11030104 | A | G | 0.041 | 0.004 | 322 103 | 5.56E-28 |
| rs3101336 | C | T | 0.033 | 0.003 | 316 872 | 2.66E-26 |
| rs7138803 | A | G | 0.032 | 0.003 | 322 092 | 8.15E-24 |
| rs10182181 | G | A | 0.031 | 0.003 | 321 759 | 8.78E-24 |
| rs3888190 | A | C | 0.031 | 0.003 | 321 930 | 3.14E-23 |
| rs1516725 | C | T | 0.045 | 0.005 | 320 644 | 1.89E-22 |
| rs12446632 | G | A | 0.040 | 0.005 | 316 758 | 1.48E-18 |
| rs2287019 | C | T | 0.036 | 0.004 | 300 921 | 4.59E-18 |
| rs3817334 | T | C | 0.026 | 0.003 | 321 959 | 5.15E-17 |
| rs2112347 | T | G | 0.026 | 0.003 | 322 019 | 6.19E-17 |
| rs12566985 | G | A | 0.024 | 0.003 | 319 282 | 3.28E-15 |
| rs3810291 | A | G | 0.028 | 0.004 | 296 261 | 4.81E-15 |
| rs7141420 | T | C | 0.024 | 0.003 | 321 970 | 1.23E-14 |
| rs13078960 | G | T | 0.030 | 0.004 | 322 135 | 1.74E-14 |
| rs10968576 | G | A | 0.025 | 0.003 | 322 061 | 6.61E-14 |
| rs17024393 | C | T | 0.066 | 0.009 | 297 874 | 7.03E-14 |
| rs657452 | A | G | 0.023 | 0.003 | 313 651 | 5.48E-13 |
| rs12429545 | A | G | 0.033 | 0.005 | 312 934 | 1.09E-12 |
| rs12286929 | G | A | 0.022 | 0.003 | 321 903 | 1.31E-12 |
| rs13107325 | T | C | 0.048 | 0.007 | 321 461 | 1.83E-12 |
| rs11165643 | T | C | 0.022 | 0.003 | 320 730 | 2.07E-12 |
| rs7903146 | C | T | 0.023 | 0.003 | 322 130 | 1.11E-11 |
| rs10132280 | C | A | 0.023 | 0.003 | 321 797 | 1.14E-11 |
| rs17405819 | T | C | 0.022 | 0.003 | 322 085 | 2.07E-11 |
| rs1016287 | T | C | 0.023 | 0.003 | 321 969 | 2.25E-11 |
| rs4256980 | G | C | 0.021 | 0.003 | 320 028 | 2.90E-11 |
| rs17094222 | C | T | 0.025 | 0.004 | 321 770 | 5.94E-11 |
| rs12401738 | A | G | 0.021 | 0.003 | 322 070 | 1.15E-10 |
| rs7599312 | G | A | 0.022 | 0.003 | 322 024 | 1.17E-10 |
| rs2365389 | C | T | 0.020 | 0.003 | 316 768 | 1.63E-10 |
| rs205262 | G | A | 0.022 | 0.004 | 315 542 | 1.75E-10 |
| rs2820292 | C | A | 0.020 | 0.003 | 321 707 | 1.83E-10 |
| rs12885454 | C | A | 0.021 | 0.003 | 320 823 | 1.94E-10 |
| rs16851483 | T | G | 0.048 | 0.008 | 233 929 | 3.55E-10 |
| rs1167827 | G | A | 0.020 | 0.003 | 306 238 | 6.33E-10 |
| rs758747 | T | C | 0.023 | 0.004 | 308 688 | 7.47E-10 |
| rs1928295 | T | C | 0.019 | 0.003 | 321 979 | 7.91E-10 |
| rs9925964 | A | G | 0.019 | 0.003 | 318 385 | 8.11E-10 |
| rs11126666 | A | G | 0.021 | 0.003 | 321 979 | 1.33E-09 |
| rs2650492 | A | G | 0.021 | 0.004 | 319 464 | 1.92E-09 |
| rs6804842 | G | A | 0.019 | 0.003 | 321 463 | 2.48E-09 |
| rs12940622 | G | A | 0.018 | 0.003 | 322 032 | 2.49E-09 |
| rs11847697 | T | C | 0.049 | 0.008 | 306 243 | 3.99E-09 |
| rs4740619 | T | C | 0.018 | 0.003 | 321 887 | 4.56E-09 |
| rs13191362 | A | G | 0.028 | 0.005 | 321 902 | 7.34E-09 |
| rs3736485 | A | G | 0.018 | 0.003 | 321 398 | 7.41E-09 |
| rs17001654 | G | C | 0.031 | 0.005 | 233 722 | 7.76E-09 |
| rs11191560 | C | T | 0.031 | 0.005 | 321 893 | 8.45E-09 |
| rs1528435 | T | C | 0.018 | 0.003 | 321 924 | 1.20E-08 |
| rs2075650 | A | G | 0.026 | 0.005 | 308 408 | 1.25E-08 |
| rs1000940 | G | A | 0.019 | 0.003 | 321 836 | 1.28E-08 |
| rs2033529 | G | A | 0.019 | 0.003 | 321 917 | 1.39E-08 |
| rs11583200 | C | T | 0.018 | 0.003 | 322 095 | 1.48E-08 |
| rs9400239 | C | T | 0.019 | 0.003 | 321 988 | 1.61E-08 |
| rs10733682 | A | G | 0.017 | 0.003 | 320 727 | 1.83E-08 |
| rs11688816 | G | A | 0.017 | 0.003 | 322 051 | 1.89E-08 |
| rs11057405 | G | A | 0.031 | 0.006 | 314 111 | 2.02E-08 |
| rs2121279 | T | C | 0.025 | 0.004 | 322 065 | 2.31E-08 |
| rs29941 | G | A | 0.018 | 0.003 | 321 970 | 2.41E-08 |
| rs11727676 | T | C | 0.036 | 0.006 | 296 401 | 2.55E-08 |
| rs3849570 | A | C | 0.019 | 0.003 | 284 339 | 2.60E-08 |
| rs6477694 | C | T | 0.017 | 0.003 | 322 048 | 2.67E-08 |
| rs7899106 | G | A | 0.040 | 0.007 | 321 770 | 2.96E-08 |
| rs2176598 | T | C | 0.020 | 0.004 | 316 848 | 2.97E-08 |
| rs2245368 | C | T | 0.032 | 0.006 | 205 675 | 3.19E-08 |
| rs17724992 | A | G | 0.019 | 0.004 | 319 588 | 3.42E-08 |
| rs7243357 | T | G | 0.022 | 0.004 | 322 107 | 3.86E-08 |
| rs1808579 | C | T | 0.017 | 0.003 | 322 032 | 4.17E-08 |

^*^BMI-increasing allele

**Supplementary Table 2:** Distribution of BMI (kg/m^2^) in strata defined by the doubly-ranked stratification method

|  |  | HUNT | | |  | UK Biobank | | |
| --- | --- | --- | --- | --- | --- | --- | --- | --- |
| Stratum number |  | Mean | Lower decile | Upper decile |  | Mean | Lower decile | Upper decile |
| 1 |  | 18.4 | 17.5 | 19.8 |  | 18.6 | 17.8 | 20.0 |
| 2 |  | 19.3 | 18.2 | 20.4 |  | 19.6 | 18.5 | 20.7 |
| 3 |  | 19.9 | 18.8 | 20.8 |  | 20.1 | 19.1 | 21.1 |
| 4 |  | 20.3 | 19.4 | 21.1 |  | 20.5 | 19.6 | 21.5 |
| 5 |  | 20.6 | 19.7 | 21.5 |  | 20.9 | 20.0 | 21.8 |
| 6 |  | 20.9 | 20.1 | 21.7 |  | 21.1 | 20.3 | 22.0 |
| 7 |  | 21.1 | 20.3 | 21.9 |  | 21.4 | 20.6 | 22.3 |
| 8 |  | 21.3 | 20.5 | 22.1 |  | 21.6 | 20.8 | 22.5 |
| 9 |  | 21.5 | 20.7 | 22.3 |  | 21.8 | 21.0 | 22.7 |
| 10 |  | 21.7 | 20.9 | 22.5 |  | 22.0 | 21.2 | 22.8 |
| 11 |  | 21.8 | 21.1 | 22.7 |  | 22.2 | 21.4 | 23.0 |
| 12 |  | 22.0 | 21.2 | 22.8 |  | 22.4 | 21.6 | 23.2 |
| 13 |  | 22.1 | 21.4 | 22.9 |  | 22.5 | 21.7 | 23.3 |
| 14 |  | 22.3 | 21.5 | 23.1 |  | 22.7 | 21.9 | 23.5 |
| 15 |  | 22.4 | 21.7 | 23.2 |  | 22.8 | 22.0 | 23.7 |
| 16 |  | 22.5 | 21.8 | 23.3 |  | 23.0 | 22.2 | 23.8 |
| 17 |  | 22.7 | 21.9 | 23.5 |  | 23.1 | 22.3 | 23.9 |
| 18 |  | 22.8 | 22.0 | 23.6 |  | 23.2 | 22.4 | 24.1 |
| 19 |  | 22.9 | 22.1 | 23.7 |  | 23.4 | 22.6 | 24.2 |
| 20 |  | 23.0 | 22.2 | 23.8 |  | 23.5 | 22.7 | 24.3 |
| 21 |  | 23.1 | 22.4 | 23.9 |  | 23.6 | 22.8 | 24.4 |
| 22 |  | 23.2 | 22.5 | 24.0 |  | 23.7 | 22.9 | 24.5 |
| 23 |  | 23.3 | 22.6 | 24.1 |  | 23.8 | 23.0 | 24.7 |
| 24 |  | 23.4 | 22.7 | 24.2 |  | 24.0 | 23.2 | 24.8 |
| 25 |  | 23.6 | 22.8 | 24.3 |  | 24.1 | 23.3 | 24.9 |
| 26 |  | 23.6 | 22.9 | 24.5 |  | 24.2 | 23.4 | 25.0 |
| 27 |  | 23.8 | 23.0 | 24.5 |  | 24.3 | 23.5 | 25.2 |
| 28 |  | 23.8 | 23.1 | 24.7 |  | 24.4 | 23.6 | 25.3 |
| 29 |  | 23.9 | 23.1 | 24.8 |  | 24.5 | 23.7 | 25.4 |
| 30 |  | 24.0 | 23.2 | 24.8 |  | 24.6 | 23.8 | 25.5 |
| 31 |  | 24.1 | 23.3 | 25.0 |  | 24.7 | 23.9 | 25.6 |
| 32 |  | 24.2 | 23.4 | 25.0 |  | 24.8 | 24.0 | 25.7 |
| 33 |  | 24.3 | 23.5 | 25.1 |  | 24.9 | 24.1 | 25.8 |
| 34 |  | 24.4 | 23.6 | 25.2 |  | 25.0 | 24.2 | 25.9 |
| 35 |  | 24.5 | 23.7 | 25.3 |  | 25.1 | 24.3 | 26.0 |
| 36 |  | 24.6 | 23.8 | 25.4 |  | 25.2 | 24.4 | 26.1 |
| 37 |  | 24.7 | 23.8 | 25.5 |  | 25.3 | 24.5 | 26.2 |
| 38 |  | 24.8 | 23.9 | 25.6 |  | 25.4 | 24.6 | 26.3 |
| 39 |  | 24.8 | 24.0 | 25.7 |  | 25.5 | 24.7 | 26.4 |
| 40 |  | 24.9 | 24.1 | 25.8 |  | 25.6 | 24.8 | 26.6 |
| 41 |  | 25.0 | 24.2 | 25.9 |  | 25.7 | 24.9 | 26.7 |
| 42 |  | 25.1 | 24.3 | 26.0 |  | 25.8 | 25.0 | 26.8 |
| 43 |  | 25.2 | 24.3 | 26.1 |  | 25.9 | 25.1 | 26.9 |
| 44 |  | 25.3 | 24.4 | 26.2 |  | 26.0 | 25.1 | 27.0 |
| 45 |  | 25.4 | 24.5 | 26.3 |  | 26.1 | 25.3 | 27.1 |
| 46 |  | 25.5 | 24.6 | 26.4 |  | 26.2 | 25.4 | 27.2 |
| 47 |  | 25.6 | 24.7 | 26.4 |  | 26.3 | 25.4 | 27.3 |
| 48 |  | 25.6 | 24.8 | 26.6 |  | 26.5 | 25.5 | 27.4 |
| 49 |  | 25.7 | 24.8 | 26.7 |  | 26.6 | 25.6 | 27.5 |
| 50 |  | 25.8 | 25.0 | 26.8 |  | 26.7 | 25.7 | 27.6 |
| 51 |  | 25.9 | 25.0 | 26.9 |  | 26.8 | 25.8 | 27.7 |
| 52 |  | 26.0 | 25.1 | 26.9 |  | 26.9 | 25.9 | 27.8 |
| 53 |  | 26.1 | 25.2 | 27.0 |  | 27.0 | 26.0 | 28.0 |
| 54 |  | 26.2 | 25.3 | 27.1 |  | 27.1 | 26.1 | 28.1 |
| 55 |  | 26.3 | 25.4 | 27.2 |  | 27.2 | 26.2 | 28.2 |
| 56 |  | 26.4 | 25.5 | 27.3 |  | 27.3 | 26.3 | 28.3 |
| 57 |  | 26.5 | 25.5 | 27.4 |  | 27.4 | 26.4 | 28.4 |
| 58 |  | 26.6 | 25.6 | 27.5 |  | 27.5 | 26.5 | 28.6 |
| 59 |  | 26.7 | 25.7 | 27.6 |  | 27.6 | 26.6 | 28.7 |
| 60 |  | 26.8 | 25.8 | 27.7 |  | 27.7 | 26.7 | 28.8 |
| 61 |  | 26.9 | 25.9 | 27.9 |  | 27.9 | 26.8 | 28.9 |
| 62 |  | 27.0 | 26.0 | 27.9 |  | 28.0 | 26.9 | 29.1 |
| 63 |  | 27.1 | 26.1 | 28.1 |  | 28.1 | 27.0 | 29.2 |
| 64 |  | 27.2 | 26.2 | 28.2 |  | 28.2 | 27.1 | 29.3 |
| 65 |  | 27.3 | 26.3 | 28.3 |  | 28.3 | 27.2 | 29.5 |
| 66 |  | 27.4 | 26.4 | 28.4 |  | 28.5 | 27.3 | 29.6 |
| 67 |  | 27.5 | 26.5 | 28.6 |  | 28.6 | 27.5 | 29.8 |
| 68 |  | 27.6 | 26.6 | 28.7 |  | 28.7 | 27.6 | 29.9 |
| 69 |  | 27.7 | 26.7 | 28.8 |  | 28.9 | 27.7 | 30.0 |
| 70 |  | 27.9 | 26.8 | 28.9 |  | 29.0 | 27.8 | 30.2 |
| 71 |  | 28.0 | 26.9 | 29.0 |  | 29.1 | 28.0 | 30.4 |
| 72 |  | 28.1 | 27.0 | 29.1 |  | 29.3 | 28.1 | 30.5 |
| 73 |  | 28.2 | 27.1 | 29.4 |  | 29.4 | 28.2 | 30.7 |
| 74 |  | 28.3 | 27.2 | 29.5 |  | 29.6 | 28.4 | 30.9 |
| 75 |  | 28.5 | 27.4 | 29.6 |  | 29.8 | 28.5 | 31.1 |
| 76 |  | 28.6 | 27.5 | 29.8 |  | 29.9 | 28.7 | 31.2 |
| 77 |  | 28.7 | 27.6 | 29.9 |  | 30.1 | 28.8 | 31.4 |
| 78 |  | 28.9 | 27.8 | 30.1 |  | 30.3 | 28.9 | 31.6 |
| 79 |  | 29.0 | 27.9 | 30.3 |  | 30.4 | 29.1 | 31.8 |
| 80 |  | 29.2 | 28.0 | 30.5 |  | 30.6 | 29.3 | 32.1 |
| 81 |  | 29.4 | 28.2 | 30.7 |  | 30.8 | 29.4 | 32.3 |
| 82 |  | 29.5 | 28.3 | 30.9 |  | 31.0 | 29.6 | 32.6 |
| 83 |  | 29.7 | 28.4 | 31.1 |  | 31.3 | 29.8 | 32.8 |
| 84 |  | 29.9 | 28.6 | 31.3 |  | 31.5 | 30.0 | 33.0 |
| 85 |  | 30.1 | 28.8 | 31.5 |  | 31.7 | 30.2 | 33.3 |
| 86 |  | 30.3 | 28.9 | 31.8 |  | 32.0 | 30.4 | 33.7 |
| 87 |  | 30.6 | 29.1 | 32.0 |  | 32.3 | 30.6 | 34.0 |
| 88 |  | 30.8 | 29.3 | 32.3 |  | 32.5 | 30.9 | 34.4 |
| 89 |  | 31.0 | 29.5 | 32.6 |  | 32.9 | 31.1 | 34.7 |
| 90 |  | 31.3 | 29.7 | 33.0 |  | 33.2 | 31.4 | 35.2 |
| 91 |  | 31.6 | 30.0 | 33.3 |  | 33.6 | 31.7 | 35.6 |
| 92 |  | 32.0 | 30.3 | 33.7 |  | 34.0 | 32.0 | 36.2 |
| 93 |  | 32.3 | 30.6 | 34.2 |  | 34.5 | 32.4 | 36.7 |
| 94 |  | 32.8 | 30.9 | 34.8 |  | 35.0 | 32.8 | 37.4 |
| 95 |  | 33.3 | 31.4 | 35.5 |  | 35.7 | 33.3 | 38.2 |
| 96 |  | 33.9 | 31.8 | 36.1 |  | 36.4 | 33.9 | 39.2 |
| 97 |  | 34.7 | 32.4 | 37.1 |  | 37.4 | 34.6 | 40.5 |
| 98 |  | 35.8 | 33.3 | 38.7 |  | 38.7 | 35.5 | 42.2 |
| 99 |  | 37.3 | 34.1 | 40.5 |  | 40.5 | 36.7 | 44.8 |
| 100 |  | 40.3 | 35.6 | 43.2 |  | 44.3 | 38.8 | 47.9 |

We report the mean, lower decile (10^th^ percentile of the distribution), and upper decile (90^th^ percentile of the distribution) of the BMI distribution for participants in each stratum created by the doubly-ranked method. In the first and 100^th^ strata, we instead report the 20^th^ and 80^th^ percentiles of the distribution.

**Supplementary Figure 1:** Associations between the genetic score and BMI in strata defined by the doubly-ranked method plotted against mean BMI in each stratum for: A) HUNT, B) UK Biobank. Error bars represent 95% confidence intervals.

A.
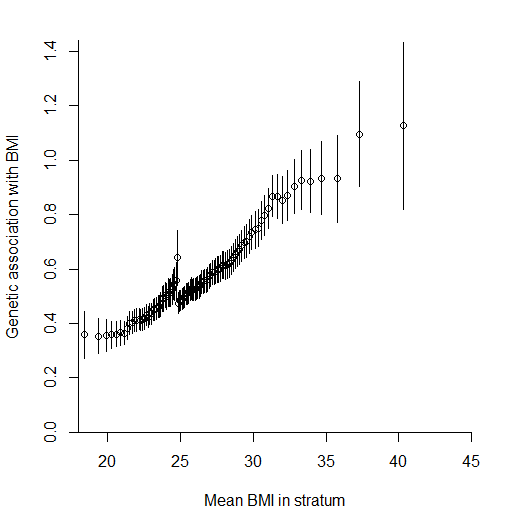

B.
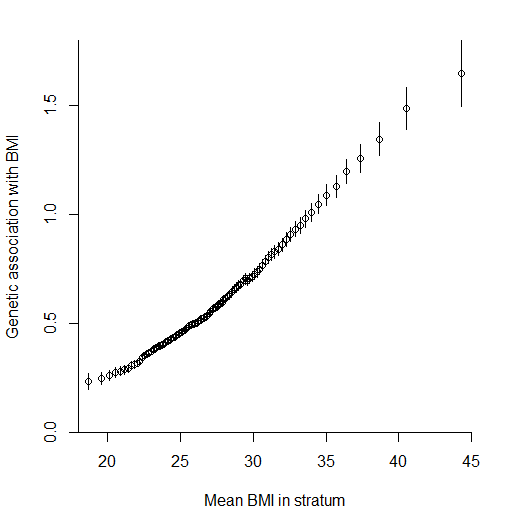

**Supplementary Figure 2:** Genetic associations with potential competing risk factors in strata of the HUNT study population

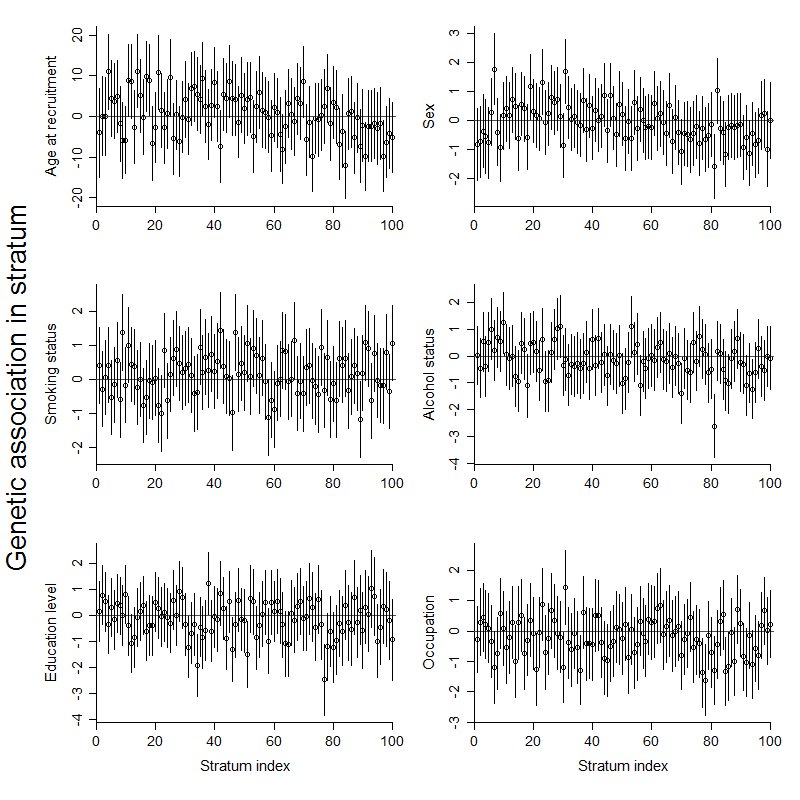

Points represent association estimates from linear (age at recruitment, years) or logistic regression (all other traits). Error bars represent 95% confidence intervals. Stratum 1 has lowest average BMI levels, stratum 100 has highest average BMI levels.

**Supplementary Figure 3:** Localized average causal effect estimates (stratum-specific estimates) for all-cause mortality plotted against mean BMI in each stratum. Estimates represent hazard ratios per 1 kg/m^2^ increase in genetically-predicted BMI. Error bars represent 95% confidence intervals.

| A) HUNT, doubly-ranked method:  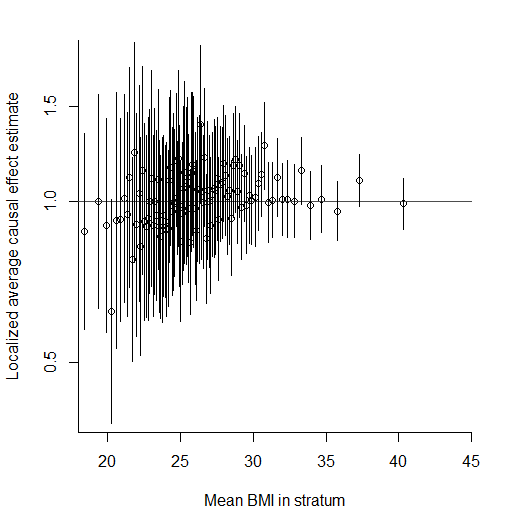 | B) UK Biobank, doubly-ranked method:  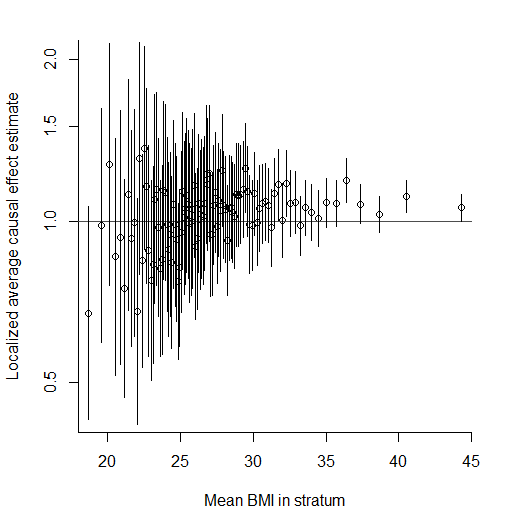 |
| --- | --- |
| C) HUNT, residual method: 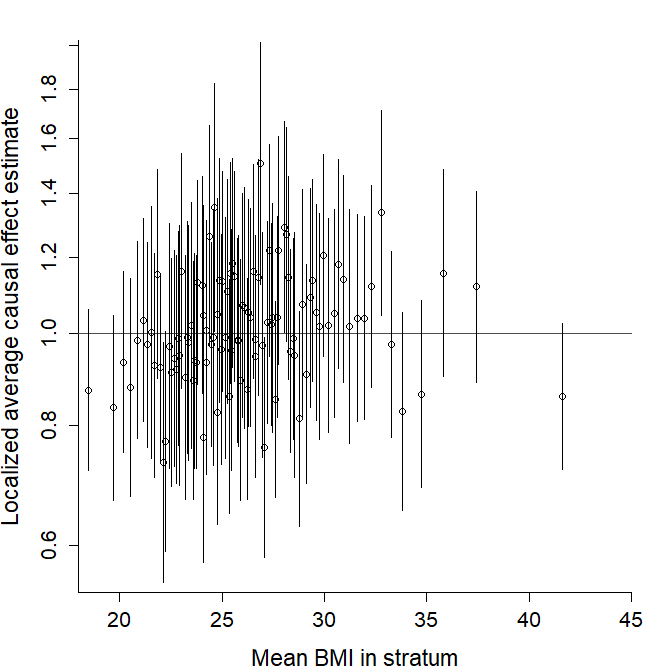 \| B) UK Biobank: | D) UK Biobank, residual method:  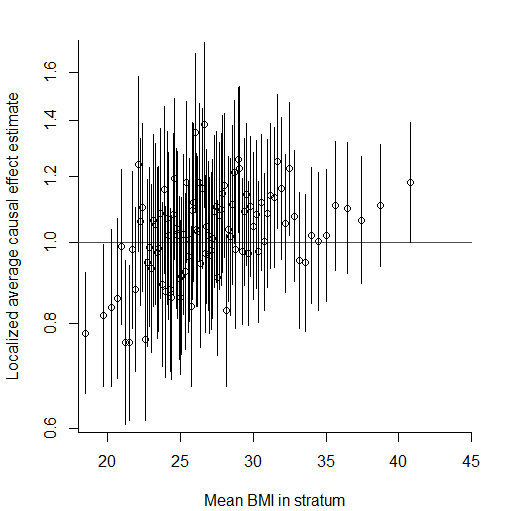 |

This presentation does not smooth over stratum-specific estimates, hence providing a representation of the causal effect of BMI which is not dependent on the choice of fractional polynomial smoothing. However, stratum-specific estimates are imprecise and variable, as they each only use 1% of the overall sample size.

**Supplementary Figure 4:** Non-linear Mendelian randomization – dose-response curve between body mass index and all-cause mortality using the doubly-ranked method with inverse-normal rank transformed BMI values in UK Biobank. Grey lines represent 95% confidence intervals. The reference value for BMI was taken as the median of the BMI distribution at 26.7 kg/m^2^.

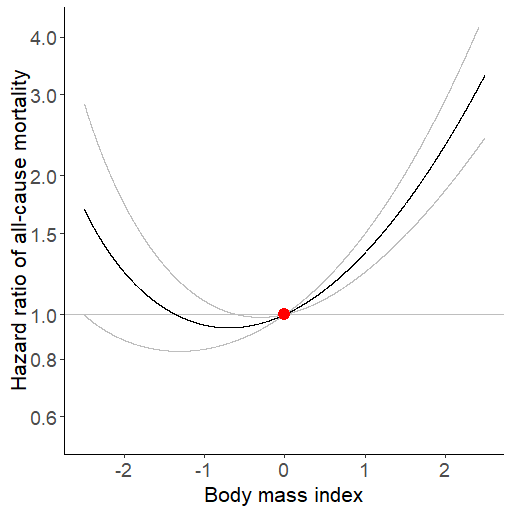

The labels on the horizontal axis represent the number of standard deviations below or above the median of the BMI distribution: for example, -1 represents one standard deviation below the median, and +2 represents two standard deviations above the median.

The reference values are:
-2 = 19.9 kg/m2
-1 = 23.0 kg/m2
0 = 26.7 kg/m2
+1 = 31.6 kg/m2
+2 = 39.2 kg/m2

**Supplementary Figure 5:** Localized average causal effect estimates (stratum-specific estimates) for all-cause mortality plotted against mean BMI in each stratum estimated in men and women separately. Estimates represent hazard ratios per 1 kg/m^2^ increase in genetically-predicted BMI. Error bars represent 95% confidence intervals.

| A) HUNT, men only:  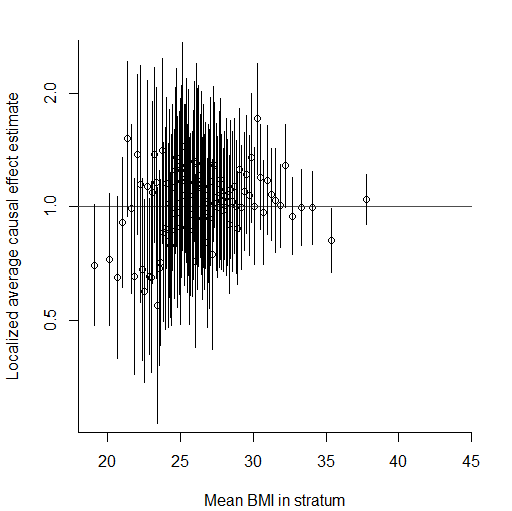 | B) HUNT, women only:  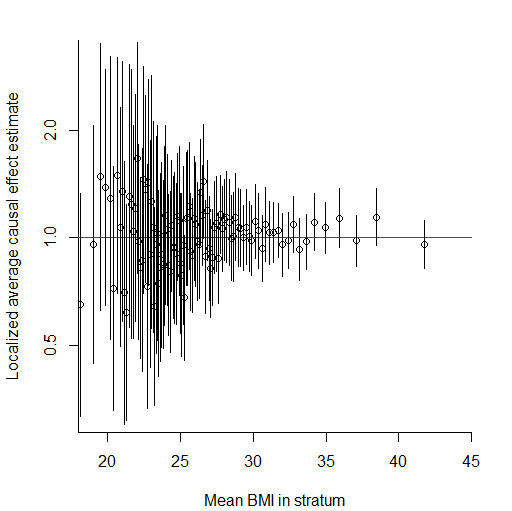 |
| --- | --- |
| C) UK Biobank, men only:  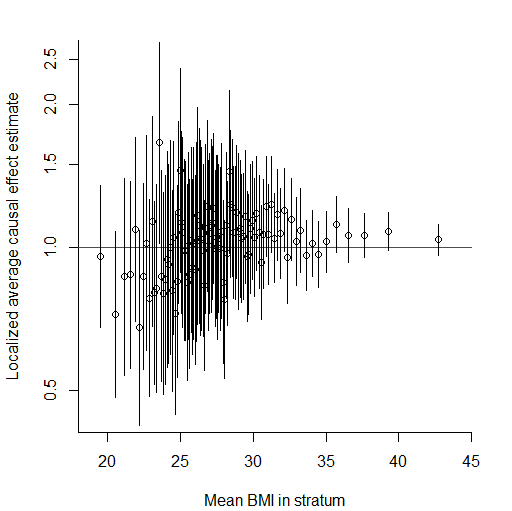 | D) UK Biobank, women only:  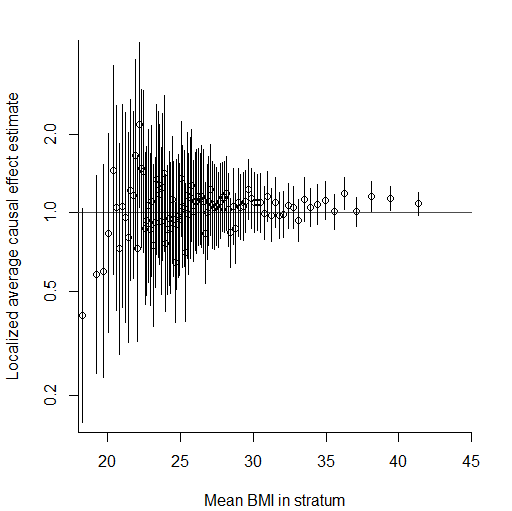 |

**Supplementary Figure 6:** Localized average causal effect estimates (stratum-specific estimates) for cause-specific mortality plotted against mean BMI in each stratum. Estimates represent hazard ratios per 1 kg/m^2^ increase in genetically-predicted BMI. Error bars represent 95% confidence intervals.

| A: Cardiovascular mortality  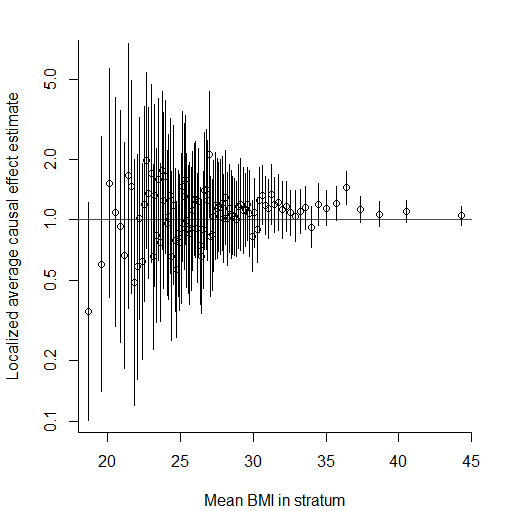 | B: Cancer mortality  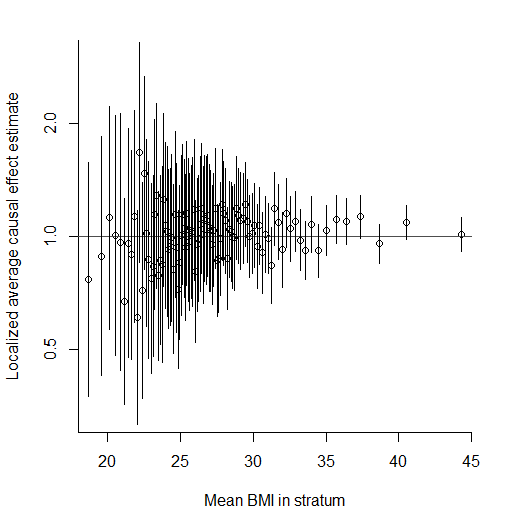 |
| --- | --- |
| C: Other (non-cardiovascular, non-cancer) mortality  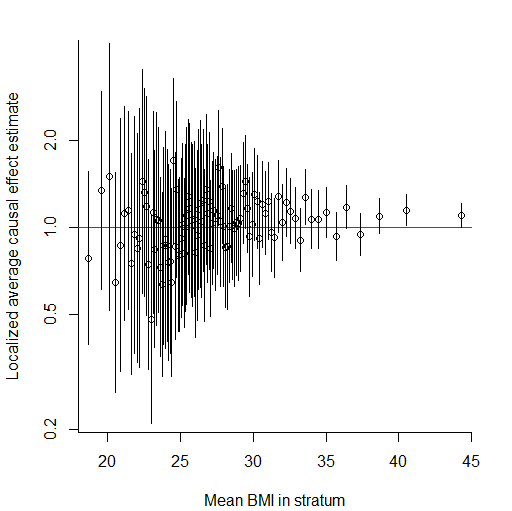 | |
